# Leprosy perceptions and health seeking behavior in Miandrivazo district, Madagascar

**DOI:** 10.1101/2025.06.10.25329324

**Authors:** Hanitriniaina Rasolofozafy, Charlotte Gryseels, Lovasoa Mbolamanana Joseph Andrianiriana, Randrianatoandro Andriamira, Epco Hasker, Bouke Catherine de Jong, Bertrand Cauchoix, Stéphanie Ramboarina, Koen Peeters Grietens, Maya Ronse

**Affiliations:** National Leprosy Program, Madagascar; Institute of Tropical Medicine, Antwerp Belgium; Fondation Raoul Follereau, Madagascar; School of Tropical Medicine and Global Health, Nagasaki University, Nagasaki, Japan

## Abstract

**Background:** Leprosy remains a severe health and social problem in several low- and middle-income countries, with two main challenges: late diagnosis, which can lead to disabilities, and the burden of stigma and social exclusion. This study explored socio-cultural factors, including community perceptions and representations of leprosy, influencing access to and uptake of biomedical care in an endemic district of Madagascar.

**Methodology:** A qualitative study using ethnographic techniques was conducted in the Miandrivazo district, southwestern of Madagascar. Theoretical sampling included patients and their families, village residents, traditional healers, health workers, and community workers. Data analysis occurred during fieldwork to discuss emerging ideas, and through thematic analysis of the full raw data.

**Principal Findings:** Leprosy was often recognized late, at the stage of visible complications such as mutilations, and perceived as mystical punishment for social transgressions. Self-medication, traditional healers, and biomedical services were avenues patients relied on when seeking care. Most initially consulted a trusted traditional healer, before turning to biomedical or other providers recommended by relatives when their health did not improve. The search of good care often required travelling to distant places at high financial cost. Care-seeking decisions were related to perceptions of the disease’s aetiology, symptoms, transmission, (perceived) treatment availability and effectiveness, and experiences or fear of stigma.

**Conclusions/Significance:** This study highlights the importance of socio-cultural factors to be considered in improving access to diagnosis and care for leprosy patients. Lack of continuous, impactful awareness about leprosy as a biomedical condition and (fear of) stigma remain major bottlenecks, within the context of a broader challenged health system.

**Author summary:** Leprosy, an infectious disease caused by *Mycobacterium leprae*, usually presents initially with benign, insensitive patches of skin with pigment loss, despite the risk of leading to irreparable nerve damage and disability if left untreated. Since the discovery of effective antibacterial drugs for leprosy, the disease has been brought under control in large parts of the world but remains a significant health problem in some countries like Madagascar. In order to interrupt transmission and prevent new patients from developing visible deformities, potential bottlenecks to early detection and treatment must be identified. Geographical and financial access to care have previously been identified as main concerns. However, this study in Madagascar revealed that perceptions of the causes and signs of the disease were main drivers of patients’ health seeking behavior. The biomedical definition of leprosy was different from its community representation as a punishment for social and moral transgressions. The disease therefore caused stigma, and the first choice of care would often be a traditional healer. The current sensitization strategy could be improved by increasing its frequency and considering the cultural understanding and stigma of leprosy in Madagascar, in order to increase early diagnosis and adherence to treatment.

## Introduction

Leprosy is a cutaneous infectious and debilitating disease caused primarily by *Mycobacterium leprae*, affecting the skin and peripheral nervous system. Generally, in the first phase of the disease, skin patches appear, then, nerve damages may follow with debilitating effects such as paralysis, claw hands and other deformities if untreated [1]. In the 1980s a combination of antibiotics (multidrug therapy or MDT) was found to be effective in controlling leprosy [2], quickly reducing individual infectiousness. MDT reduced the global burden at the end of the 2000s when the elimination of leprosy as a public health problem was declared at global level; 107 countries had reached prevalence rates of less than 1 case per 10,000 inhabitants [3].

However, two decades later, in 2023, only 53 countries recorded no cases. Madagascar is one of the 23 countries that are prioritized for leprosy control by the WHO [4]. Despite active case finding campaigns in certain endemic districts in accordance with (and even preceding) the WHO guideline for preventing disabilities in new leprosy cases by early diagnosis and treatment of cases, in 2023, 1659 new cases were detected in Madagascar, 18% of whom were already with advanced disability at the time of diagnosis, and 6% were children under the age of 15 [5,6].

In its Global Strategy 2016-2020, the WHO recommended single-dose prophylaxis with rifampicin for contacts of leprosy patients as a study in Bangladesh reported a 57% reduction in risk of leprosy [7]. The WHO recommends countries to study how the implementation of rifampicin chemoprophylaxis can be adapted to their contexts. Indeed, contextual factors such as distance to healthcare facilities and financial barriers are often mentioned as factors impeding access to and the completion of treatment for leprosy [8] [9]. Stigmatization is also reported to lead patients to conceal their disease and not seek care [10–13]. It can also lead to psychological stress and lack of income for patients and their families, leading to further delays in treatment. Lack of knowledge about the warning signs of the disease can further jeopardize seeking timely care [14].

Malagasy proverbs, expressions and adages [15], however, show the local burden, such as *“mitoka-monina ohatrin’ny boka”* (“to isolate oneself like a leper”) when a person is reproached for not getting together with others; or, *“boka misaka foza ka manosika azy hilentika”* (“a leper who wants to look for the crab in the mud will sink in it even further”) referring to the harsh reality of leprosy limb amputations. Disease perceptions, as is also the case for leprosy in Madagascar, are embedded in cultural logics [16], and insights into those logics are key to fight leprosy.

This research was part of the clinical trial *Post Exposure Prophylaxis of Leprosy (PEOPLE)* on the administration of rifampicin as post-exposure prophylaxis to contacts of leprosy cases in Madagascar and the Comoros (ClinicalTrials.gov Identifier: NCT03662022). This embedded qualitative study explored the socio-cultural factors influencing the effectiveness of trial-related interventions, including community perceptions and representations of leprosy.

## Methods

### Study context: the PEOPLE clinical trial

The PEOPLE trial mainly aimed at determining an optimal implementation scenario for prophylaxis in contacts of leprosy cases with the antibiotic rifampicin [17]. The trial implemented door-to-door screening for leprosy by a team of health workers, different types of biological sampling, and distribution of rifampicin to contacts in three different ways depending on the study arm. Prior to implementation, sensitization sessions were conducted in the selected villages by community health workers to convey messages about the door-to-door screening activity and about leprosy in general. All leprosy patients identified were offered free treatment in accordance with the national program protocol (see [18] for more details). This anthropological study was carried out ancillary to the clinical trial to explore implementation bottlenecks. Our qualitative research further explored the socio-cultural factors that influenced the implementation of the clinical trial, including community perceptions of the disease and healthcare utilization.

### Study sites and population

This qualitative study was carried out in nine out of the 20 trial *fokontany* (the administrative subdivision below the commune, equivalent to a village) of three rural communes in the Miandrivazo health district in south-west part of Madagascar. For confidentiality purposes and potential stigmatization concerns, the concrete study villages are not mentioned by name. There are several ethnic groups living in the study sites which can be divided into two groups: those known as native groups, i.e. the Bara and Sakalava ethnic groups; and other groups such as the Merina, Betsileo (from the highlands of Madagascar), Antanosy and Antandroy (from the south), and the Antemoro and Korao (from the south-east). In Madagascar, the health system follows a pyramidal structure with at the top, the most specialized care being provided in 23 hospitals in nine cities including the capital. The referral system supposes patients get referred from lower and more general care structures available in communes or *fokontany* to more specialized hospitals in districts or regions. At the lowest level of the pyramid primary health care centers (called CSB) are envisioned as the first point of contact of the population with the formal health system. The healthcare centers, which are mainly run by nurses, are meant to be available in villages (but nearly 66 % of villages are more than 5 km from a health center [19]). When we mention health centers in this manuscript, we refer to these types of health structures. In the area of the trial study sites, four health centers exist. For leprosy, it is expected that patients can consult there for diagnosis and receive free treatment and follow-up. However, once treatment is initiated, community health workers residing in *fokontany* may be in charge of following-up patients, including the provision of their monthly medication. It is worth noting that apart from the treatment of leprosy, the treatment of most other skin conditions, even the common and minor ones, is not free of charge in Madagascar and often unavailable in health centers.

### Sampling

We proceeded by theoretical and snowball sampling [20], with concomitant data collection and analysis. Emerging theories determined the next piece of information to be collected and (key) informants involved. The sampling techniques were used to include different types of informants, including people affected and not affected by leprosy; healthcare providers (community workers, public and private health center staff, clinical trial staff, traditional healers); and local authority figures.

### Data collection

Fieldwork was carried out by a team consisting of a female anthropologist affiliated to the National Leprosy Program of Madagascar and one from the Institute of Tropical Medicine in Antwerp Belgium. In the field, gatekeepers were meticulously selected to facilitate movement through the villages and approach key informants. Frequent field visits, including overnight stays, were part of our approach to building trust with communities and immersion in the study context. During each field visit, the anthropologist(s) spent several days in the villages at each of the rural study sites. These data-gathering periods were interspersed with intermittent analyses to guide the further sampling and themes to be explored in greater depth. Semi-structured and in-depth interviews (IDI), group discussions (GD) and participant observation (PO) including informal conversations (IC) were carried out intermittently between September 2019 and December 2020. The interviews were conducted in Malagasy and sometimes in French when addressing the local medical study team, health workers or teachers in the village. Through interviews with key-informants among political, administrative, religious, and traditional authorities, we were able to select gatekeepers that were residents of the villages, had a good reputation and were well known and trusted by community members. We usually conducted PO, including informal conversations, at the beginning of the data collection to gain main ideas about the topics, to select potential informants for in-depth interviews through an IDI or GD and when sensitive topics were intended to be broached or when participants refused an official interview.

### Analysis

The PASS-model [21], a model founded on theory to guide the formulation of questions on health seeking behavior and access to care, helped in structuring initial question guides, thematic analysis and data interpretation. The analysis was carried out in two phases during the study. First, analysis was intermittent between the data collection periods (cf. theoretical sampling explained above), followed by internal discussion between team members to guide the next steps of data collection and go into greater analytic depth. Secondly, raw data was processed in its textual form, translated in French (for the data collected in Malagasy) and thematically coded to generate analytical categories using Dedoose qualitative data processing software [22]. The coded excerpts were then compared and interpreted to identify emerging themes and summarize findings.

### Ethical considerations

The research was approved by the Biomedical Research Ethics Committee of the Madagascar Ministry of Public Health (reference number: 032/MSANP/SG/-AGME /CNPV/CERBM) and by the Institutional Review Board of the Antwerp Institute of Tropical Medicine (reference number: 1255/18). Verbal informed consent was obtained for all participants in this study as approved by both ethical review boards. Verbal consent allowed to reduce the formality of the procedure, allowing a more natural flow of discussion. It is important to build trust between researcher and participant and, thus, to ensure the quality of data collection, especially when applying informal techniques such as informal conversations [23] The research team presented the rationale for their presence in the villages and the information about the work to be carried out, after which consent was sought from communities leaders and participants before data collection began. Confidentiality aspects were discussed in depth and time was taken for potential questions. For minors (under 18), consent from parents was sought and ascent from the minor. We chose the place to have the conversation according to both parents’ and the minors’ preferences. All information that could lead to the identification of respondents or their villages has been removed from this publication.

## Results

### Study Participants

We conducted 65 in-depth interviews; 10 group discussions; 39 informal conversations and 57 observations. See Table 1. Participants were men and women from different age categories: children, young people, young adults, adults, elderly people. Of the 20 patients included in our data, six were enrolled as patients in the PEOPLE trial. In most cases, we obtained data directly from patients (18 individuals), but sometimes from family members (two individuals). The main socio-economic activities of the informants included farming and fishing, with other varieties such as healthcare work (biomedical and traditional), business (trading, selling), handicrafts making, brick making, teaching, driving and panning for gold in rivers. There were other participants of various social statuses: *fokontany* chiefs, village committee chiefs and community workers.

**Table 1.** Distribution of participants by individual characteristics*.

|  |  | IN-DEPTH<br>INTERVIEWS<br>(n=65) | PARTICIPANT<br>OBSERVATION<br>(n=57) | INFORMAL<br>CONVERSATIONS<br>(n=39) | GROUP<br>DISCUSSIONS<br>(n=10) |
| --- | --- | --- | --- | --- | --- |
| GENDER | Mixed | 17 | 47 | 13 | 02 |
|  | Female | 27 | 05 | 13 | 03 |
|  | Male | 21 | 05 | 13 | 05 |
| AGE | Mixed | 09 | 38 | 10 | 05 |
|  | <18 | 01 | 01 | 00 | 00 |
|  | [18-25[ | 07 | 02 | 00 | 01 |
|  | [25-50[ | 22 | 13 | 15 | 02 |
|  | [50-60[ | 18 | 03 | 13 | 02 |
|  | ≥60 | 08 | 00 | 01 | 00 |
| PROFESSION | Farmer | 53 | 33 | 21 | 08 |
|  | Fisher | 05 | 00 | 01 | 00 |
|  | Health professional | 02 | 11 | 08 | 00 |
|  | Community (health) worker | 01 | 00 | 02 | 00 |
|  | Traditional healer | 02 | 00 | 00 | 00 |
|  | Teacher | 01 | 01 | 02 | 02 |
|  | Mixed /other | 01 | 12 | 05 | 00 |
| LEPROSY | Leprosy affected and/or relatives | 13 | 06 | 08 | 01 |

\* One informant can be included in more than one type of data collection

### (Non) perceived signs and severity of leprosy

Leprosy was commonly referred to as *habokana (the fact of having leprosy)*. The illness was identified by community members when the signs suggestive of leprosy were visible: sores and ulcers on the skin, amputations of limbs, facial deformities, and sometimes reactions with neurological or systemic complications. Therefore, leprosy was also known in the community as *manapatapaka*, meaning the disease that amputates; and, by older participants as *aretin-koditra*, i.e. skin disease.

Some patients diagnosed in the early stages of their disease with only skin patches stated they were surprised when receiving the diagnosis, indicating that less pronounced/visible signs were not always interpreted as leprosy.

Skin patches could be embarrassing and even be perceived as a sign of disease, but not necessarily leprosy. The wife of a leprosy patient who lived in a village with a health center said that she had noticed the (insensitive) skin patch on her husband’s knee, the sign that led to his diagnosis for leprosy by the team of the project, for about nine years without this prompting a search for care. “*On his knee, there is a patch that doesn’t itch, is not painful but it was there since I saw it the first time when we got married*” (Female participant, adult, shopkeeper, IDI).

Moreover, skin patches were often perceived to be caused by frequent contact with river or lake water, common in children and healing spontaneously in adulthood: “*We don’t know how, but the spots disappear in adulthood“* (Female participant, adult, farmer, GD). These skin problems were interpreted as non-severe and therefore did not lead to an active recourse to care. This perception is in contrast with the representation of leprosy, which is perceived as dangerous, and associated with severe, advanced signs of the disease.

The less visible neurological complications such as paresthesia, or weakness of a limb, and even clawed fingers, were also not clearly identified as signs of leprosy. When these signs appeared, other explanations were considered more likely causes of the symptoms than leprosy, such as long-lasting wounds, curses cast by adversaries, stings from fish fins, etc.

### Perceived aetiology of leprosy

The presence of skin patches did not automatically prompt people to seek care. Certain illnesses were perceived to fall within the domain of the healer’s capacity to treat, while other illnesses, such as suspected malaria or stomach-ache, should be taken to a medical doctor. “*There are different categories of illnesses […] When your illness is one that should be treated at the healer, but you take it to the doctor, you won’t be cured.” (Male patient, adult, fisher and fishnet maker, IDI)*. In case of perceived signs of leprosy, their aetiology was primarily considered mystical in nature.

#### Leprosy as a consequence of social tensions and moral transgressions

Leprosy was mainly considered as a curse or punishment for immoral and antisocial behavior. Spirits of the land (*angatry ny tany*) were perceived to inflict the disease on thieves or on those who had been caught by the *kialo* or a means of protection against theft, provided by traditional healers (“ombiasy”), that is placed in the field so that anyone who touches the crop, both thieves and people passing through the fields, will prompt the spirits to inflict a disease such as leprosy.

Similarly, respondents reported that leprosy could be the consequence of a curse. Following a disagreement within the community, people could punish each other by putting an *ody* (object used to cast a spell) in contact with their adversaries by various means, like the exchange of money, or work tools such as a spade.

An additional perceived cause was the infringement of cultural taboos, such as eating mutton or pork; marrying despite parents’ opposition; touching and wearing gold (depending on the ethnicity); eating hens or even buying clothes with money obtained from their sale. When leprosy was thought to be caused by breaking a taboo, it was also thought to be incurable: “*No, I have never seen a person cured of a skin disease [leprosy]. It never cures*” (Female participant, older adult, farmer (but now retired), GD). This notion of incurability was expressed above all by older informants for whom respecting taboos was considered most important.

#### Leprosy as latent processes

Leprosy could belong to the local illness category *aretina an-tsoson-koditra* (i.e. disease inside the skin). These illnesses were thought to develop inside the body, persist in the patient over time and at some point, express themselves externally through symptoms. The disease could manifest for several reasons, including the consumption of certain foods – “*It comes from tasting something you couldn’t tolerate, and it appears on the outside*” (Female participant, adult, farmer, IDI) – or the prolonged persistence of disease – “*It itched, and I scratched, and it left patches. People who look at it call it leprosy, but it’s an[other] illness that lasted and then turned into leprosy”* (Female patient, adult, farmer, IDI). When the disease manifested itself, it was perceived that it could progress from a milder to a more serious form if left untreated or even turn into a new illness. As a participant voiced: “*For illnesses, for example, you have a stomach-ache, […] when you don’t take the medication for stomach-aches, it leads to other illnesses*”. (Male patient, adult, fisher and fishnet maker, IDI).

#### Leprosy as a biomedical condition

The notion of leprosy as an infectious disease caused by micro-organisms was not widely held within the community. This knowledge seemed to be acquired only in the event of prolonged contact with the biomedical care system, as in the case of patients who had benefited from care in leprosy complication management centers, where they could stay several months.

### Perceived transmission of leprosy

Some informants reported that leprosy is transmitted from one generation to the next. This perception was also accepted by patients where the illness clustered in the family and contributed to the gravity associated to the disease: “*The doctors say it’s transmissible, but what scares me is that it could be transmitted to my child*” (Male patient, adult, farmer, IDI). However, patients that were the only cases in their family often expressed that the disease was not transmissible. A patient said: “*My disease is not transmissible. If it was transmissible, my kids would all have been infected by now*” (Female patient, adult, farmer, IDI). People without leprosy often reported to prevent transmission by avoiding contact with patients. Risk for transmission was seen when having close contact (*mifanosonosona*), bathing in the same water, eating or sleeping with a patient or touching their personal effects such as clothes or eating utensils, touching a sick person’s corpse on the leaking parts of his or her body.

### Treatment options for leprosy

#### Home treatment

According to respondents, certain types of plants could be used to treat leprosy symptoms. The bark of a tree known as “bokabe” (or great leper, so called because of its rough appearance) was said to have the ability to make skin spots invisible when rubbed vigorously over the affected skin. Other kinds of home treatments included other types of pieces of wood; a solution of boiled leaves applied to the affected skin; and crushed leaves as poultice for skin treatment in an advanced stage of the disease. Other self-medication included a mixture of honey and fine rice bran on the wounds covering them and keeping them warm to improve the healing process. A mixture of oil and table salt for dry or itchy skin after the end of treatment was also mentioned by healed patients.

#### Traditional healing

Patients often reported that they first visited traditional healers to explain the nature and origin of their symptoms. Causes identified by traditional healers were often either curses or transgressions of taboos or social norms. When asked about the origin of their illness, patients often described the cause provided by the healer, demonstrating their trust in healers, even in instances where healthcare workers were simultaneously consulted.

Traditional healers would identify those responsible for the patients’ misfortune through divination and provide treatments that resembled self-medication options like leaves and bark to be boiled, and the vapor inhaled, or used in solutions on the affected parts of the body or to drink.

#### Biomedical treatment

In the study setting, patients could seek medical treatment at biomedical public, private, or confessional health centers (managed by religious). While four health centers existed in the study area, this still meant that many people needed to spend a considerable amount of time to get there. Biomedical care for leprosy mainly consists of MDT (multi-drug therapy) to be taken for six- or twelve-months contingent on the paucibacillary or multibacillary form of disease. Patients described the efficiency of the treatment: “*When you take medicines, the strength of the disease is dissolved”* (Male patient, adult, farmer, IDI). Subsequently, there were specialized centers for the management of neurological complications or wounds on the limbs, which were often confessional. These confessional centers were located either in the same district, or in a different one, ranging from 40 to more than 200 kilometers from the patients’ villages. Patients expressed high levels of appreciation for these confessional facilities, citing the attentive care and accommodation provided as key factors. Apart from the MDT, some patients, in particular those who had been in a center for complications management, said they needed other medicines for specific symptoms such as pain in the limbs, which referred to the corticoid treatment: “*These medicines [showing the blister packs of prednisolone] are what accompany the other treatment [the multidrug therapy]”* (Male patient, adult, farmer, IDI).

The majority of patients had consulted with a health center at some point during their itinerary of care for leprosy.

### Factors driving choice of care for leprosy

#### Preference for traditional care system

When visible and advanced signs appeared, patients usually sought care with traditional healers first. The representation of leprosy as a potential punishment led patients to search for the cause and to repair it. In that respect traditional healers reassured patients and satisfied their expectations: to get an answer on what is wrong with them, why it has happened and how to cure it. Self-medication was another option, but only when patients considered the aetiology of the symptoms not to be associated with punishment and/or it could be used concomitantly with other types of care (e.g. traditional and/or biomedical). If a consultation with a healer did not result in the cure the patient had hoped for, the patient and his/her relatives would opt to seek the services of other healers. However, when successive treatments by healers could not cure the disease, they would often turn to biomedical options such as health centers. Patients in our study often sought care at health centers as a last resort. Only one single patient reported directly seeking care at a health center as a first step; this individual had resided in an urban setting throughout their childhood and adolescence and was used to going to biomedical care services in the event of illness.

#### Biomedical care provider attitudes and practices

Care-seeking decisions could also change after the experience gained in one chosen care option. To illustrate, one woman decided to go to a healthcare worker in her village when she was unable to recover after consulting traditional healers, but she felt unwelcome because the healthcare worker made remarks about her clothes not being clean. She had to unsuccessfully visit another health worker in the village before being properly referred to a confessional leprosy center in the district by a community health worker, about 40 kilometers from her village, where she went and finally received the diagnosis of leprosy with the corresponding care and extensive support (nutrition, clothes).

In many cases, patients had consulted with more than one health center before being diagnosed with leprosy and initiated treatment. The time between the first and last biomedical encounter prior to diagnosis ranged from several months to years. This indicates the possible lack of knowledge about the disease among healthcare workers, which may result in missed diagnoses.

The way in which the health worker communicated the diagnosis also played an important role in the acceptance of both diagnosis and treatment by patients. According to health workers working with leprosy patients, the expression “having leprosy” was more readily accepted by patients as it refers to being affected by any pathology that is supposed to be temporary. Conversely, using the adjective “being a leper”, implied a permanent condition with irreversible disabilities such as amputations and deformities and was linked to stigma. A door-to-door screening team member who tried different strategies to reach out to patients in communities said, “*When you ask a patient what their disease is, they say ‘leprosy’, but when you later say to them, ‘you are a leper’, they reply, ‘no, I am not a leper’*” (female health professional, adult, PO). Health workers often announced the diagnosis of leprosy by using the term “skin disease” and avoided direct terms referring to leprosy so as not to shock patients.

#### Accessibility and affordability of care

Affected people sometimes visited multiple remote villages, at several hours’ walk from their residence in search of the care they thought necessary to cure their illness, such as a good healer, known or recommended by relatives. In some cases, the financial burden associated with this pursuit could be significant, with some families spending considerable sums of money and sometimes resorting to selling their cows to seek treatment from a certain health provider. “*At that time, we were still well off, the zebus had not yet been taken away by the thieves, and we were able to get treatment from the doctor and various healers. I can’t remember where we didn’t go to search for care”* (Male relative of patient, adult, farmer, GD). Many respondents were unaware of the availability of free leprosy treatment before the screening team came to the study sites.

### Factors driving treatment adherence

#### Knowledge of treatment regimen

Even when patients initiated biomedical treatment, there were reasons for discontinuing treatment that could be related to the lack of understanding about how to properly take the medication. A lack of clarity about how long and how often to take the medication led some patients to stop their treatment before the end of the prescribed course and/or take it incorrectly. For example, one patient explained how he had finished his MDT strip (which is supposed to be taken over the span of one month) within a week and two days, then did this a second time and stopped the treatment because there was no treatment available at the dispensary and he felt cured.

> *“For me, it’s not every month [that I go to the doctor for the medicine] but every time the medicine runs out. In the morning, I swallow two [capsules], and in the evening, I also swallow two. And it’s all over in a week and I go and get some more. I go there [to the dispensary] and there’s no more medicine, and by the end I’m demotivated” (Male, adult, patient, farmer, IDI).*

The inability to comply with medical guidelines was also mentioned as a reason to abandon treatment. One patient, for instance, stopped his treatment because health workers told him that he could not drink alcohol while on treatment and he could not stop drinking alcohol.

#### Reported side-effects of the treatment

As the following quotes illustrate, malaise, hot flushes, a feeling of weakness and headache were mentioned as side-effects which led some patients to interrupt their treatment (MDT), sometimes taken inappropriately:

> *“When I take this medicine, I feel weak, I go to work and I can’t work, I’m all tired […]. That’s what it does. You get like drunk when you take this medicine.”* (Male, adult, patient, farmer, IDI). This patient had daily taken more tablets than prescribed.

> *“[…] [I]t [the medication] gave me a headache, so I stopped. I’ve only been taking this drug for three days and I’ve felt discomfort in my body and at the end I told them [the screening team] that I can’t stand this drug. ‘If there’s another one’, I said, ‘I’ll take it, but if it’s the same drug, I can’t take it’.”* (Male, adult, patient, farmer, IDI).

#### Diagnostic perceptions

Furthermore, in some cases, patients shared how they refused the treatment for leprosy. This happened mostly when the diagnosis was made during active screening in the community, and the illness was diagnosed at an early stage. In this situation, some patients rejected the diagnosis as they felt no specific discomfort or symptoms that seemed serious in relation to the disease.

### Social impact of stigmatization

Some patients chose to seek care far away from their village to avoid stigmatization. As leprosy was often interpreted as an illness of people who have committed theft and other moral and social transgressions, leprosy was seen as a shameful disease that led to stigma. In addition, the fear of contagion contributed to stigmatization.

The stigma towards leprosy patients, although attenuated in its manifestations over time according to community members, persisted but seemed to differ between contexts. In some villages, patients were excluded from everyday life in the community, even within a household. In a few cases, patients slept in a separate room annexed to the house while most of the time, families we visited lived in a house with a single room. Sometimes, patients ate separately from their family, with separate plates and utensils while the rest of the family would share their food: “*If they’ve set aside my plate, it’s because people are afraid of it [the disease]. But if they were not afraid of it, they wouldn’t put my plate aside. If I cook separately, it’s because people are afraid”* (Male patient, adult, wood seller, IDI).

Informants indicated that a divorce could ensue in the event of a diagnosis of leprosy, and that marriages to patients or their family members can be avoided or cancelled on ground of the diagnosis. The reason for this social distance remained the fear of transmission. Despite a few patients where we observed or were told that social relationships were (self-)limited due to the illness, in many cases, the patients in our study were living with their families, were married and/or had children, and mentioned having family, colleagues and/or friends with whom they had regular contact.

With regard to the relationship between the community members and the healthcare staff, some patients feared that the care provided by the medical teams visiting the villages throughout the screening process might exacerbate the stigma they were already facing. This was linked by some patients and their families to the fact that on the one hand, community health workers and doctors talked about the disease by saying that it is transmissible through contact, which accentuated stigma. On the other hand, a lack of trust in health workers would also have arisen due to the fact that some patients suspected healthcare workers of disclosing their illness. “*We’re stunned, it’s the doctors who are behind this [disclosing our illnesses], they said this thing [my disease] is transmissible. […] Nobody knew about my illness, but it was the people who examined me who spread the word […].*” (Male patient, adult, farmer, IDI)

## Discussion

In the context of Madagascar, one of the 23 countries prioritized for leprosy control by the WHO, the substantial delay in seeking and receiving care for leprosy, as shown in our study, constitutes an alarming problem. Around one fifth of new leprosy cases in Madagascar present visible deformities at the time of diagnosis [5]. Our research shows how geographical, socio-cultural factors and stigma, present obstacles to the accessibility of appropriate care.

### Biomedical definitions of leprosy at odds with local socio-cultural interpretations of leprosy symptoms

Seeking timely care based on the recognition of the warning signs of leprosy (i.e. passive case detection) constitutes the main programmatic approach for preventing invalidities and further transmission in the community. However, in Madagascar leprosy patients are often seen in the formal health system only at later stages of disease progression, namely with visible deformities and complications with neuritis. Our research demonstrates that community perceptions of leprosy do not include early warning signs such as skin patches and milder symptoms of leprosy, impeding timely seeking of a diagnosis and treatment. Indeed, early-stage disease and milder symptoms of leprosy are often not interpreted as leprosy, nor any other severe disease, and consequently do not lead to seeking care.

In this study in Madagascar, the perception of leprosy is linked to immoral and antisocial behavior which can have effects on stigma and access to care, and contribute to the notion that leprosy is incurable according to some members of the community. A study of a community in Nigeria showed that leprosy is associated with dirtiness and immoral behavior and is therefore considered as a dishonorable disease [24]. This perception accentuated the stigma and further limited patients’ access to care in health centers. Curing leprosy is not as obvious as in other diseases, since it can lead to irreversible disabilities. Even long after treatment, leprosy reactions can occur, which throw cured patients back into a state of serious illness, and lead to visible and irreparable damage if left untreated. In the past, when the disease was not yet treatable, the image that prevailed was that of patients who were isolated from the community to prevent transmission, and who carried disfiguring disabilities until their death. Decades later, this image has not yet disappeared from the collective representation, especially for older participants. Madagascar is, however, no exception. In many places of the world where leprosy has existed, including Europe, perceptions of leprosy have remained associated to a disfiguring life-long condition. In Japan, for instance, until as recent as 1997, discriminatory laws were still in place [25] and (official) isolation, sterilization and abortion were practiced even though MDT was already widespread [26,27], illustrating that perceptions have been ingrained since ancient times and continue to exist [28–30].

### Bridging the gap between the biomedical and traditional care systems

According to our results, patients were more likely to go first to a traditional healer than to a formal (biomedical) healthcare center or practitioner to be treated for perceived (and frequently already advanced) signs of leprosy. In the case of symptoms that were interpreted as more serious, patients would sometimes visit villages several hours’ walk away from their residence in search of a good healer, and answers about their illness. Distance, although a real difficulty, was not therefore a major bottleneck in achieving access to healthcare for leprosy, insofar as patients were able to overcome it when feeling the need to seek care. Depending on the results of the healers’ visits and often with the onset of complications, the patient would turn to biomedical care as a last resort. While the focus of our study was on leprosy, our findings on health seeking practices are likely to apply to other health issues as they are embedded in broader logic and perceptions of health and illness, and in similar challenges relating to access to care.

Several studies in Asia and Africa show that for leprosy [10,13,14,31] (as with other health issues, see e.g. [32–34]), traditional healers are the first option of seeking care before attending biomedical healthcare facilities. Opting to first consult with healers is suggested to be a factor of case detection delay [10,13,14]. Integrating or collaborating with healers in formal healthcare has been practiced in many contexts and health topics [32–34]. Involving them as social workers to refer patients for timely consultation in health centers could be a strategy to induce both early detection [35] and adherence to treatment. In a study in Indonesia, healers’ activities were valorized as they received training on leprosy diagnosis and treatment using a leprosy booklet helping them during consultations, and then their practices were evaluated [36]. According to a study in Benin, multiple visits to a healer constituted a factor in delaying treatment and resulted in patients being seen at an advanced stage of the disease, yet health workers’ lack of skills in recognizing the early signs of leprosy was also a factor in the high proportion of patients with grade 2 disability [12]. These findings are consistent with those of our study and highlight the need to consider and empower all stakeholders who might be involved in leprosy control for an integrated approach.

For the communities in the study sites, lack of awareness of the possibility of free treatment was also a barrier to obtaining biomedical care. In addition, sometimes, the diagnosis of leprosy would not be made at the first visit to a health center, and in some cases, patients had to consult two or three care providers before receiving a diagnosis and appropriate treatment. This highlights the lack of knowledge among some health workers about leprosy, but also the unsatisfactory welcoming of patients by health workers.

A delay in diagnosis has also been mentioned in other studies, such as in China for instance, where patients could only be treated after a delay of up to 202 months and after visits to different health centers with different specialties [37]. Other presented findings resonate with additional work conducted in different settings. For instance, in a study carried out in Ethiopia, it was revealed that knowledge of leprosy is lacking for patients and leads to a delay in diagnosis and treatment [31]. According to another study in three states of India, when questioned about the reasons that prevented them from seeking care, the patients mainly mentioned that they did not know that these skin patches corresponded to an illness, or they thought that it could disappear spontaneously. Only a small proportion of the study population mentioned not having the financial means to access a health center and four to 51 % did not know of a health center where they could go for treatment. [38].

### Overcoming (fear of) stigma and increasing awareness of leprosy as a biomedical condition

The representation of leprosy transmission as being hereditary and inter-human, and the perception that it can result from a punishment, fed the stigmatization and led some patients to deny and hide their disease. Avoiding biomedical care was a way not to reveal their illness. Some patients moreover expressed a mistrust toward healthcare workers with regard to respecting the confidentiality of their illness.

Our findings converge with those from Feenstra et al. in Bangladesh, which illustrate the dilemma that patients may face in such a situation between the necessity to disclose the diagnosis as a way to raise awareness and protect family members from infection, and the fear of stigma that could result from being identified as affected by leprosy [39].

Our study additionally highlights the need to improve general awareness about leprosy, both in terms of the content of health education messages and of the way they are delivered, since knowledge about leprosy, its transmission patterns and treatment options was not optimal, and stigma still manifested in various ways. While intensifying awareness by increasing the number of districts to be targeted for leprosy sensitization coupled with the reiteration of sensitization within these communities, would undoubtedly constitute a commendable approach, a crucial and cost-effective recommendation would be the re-evaluation of the content of awareness-raising messages. This should incorporate the early signs of the disease to promote early diagnosis, and caution against the risk of severe outcomes if left untreated, in addition to the fact that from the day a leprosy patient starts treatment, infectiousness is almost zero [40]. In recent years, the national program has started using visual aids depicting skin patches as early signs of leprosy and this initiative seems to start producing positive results. Additionally, our study shows that patients need to be provided with a greater amount of information about (free) treatment options and providers, and the need to be informed about leprosy diagnosis in a socially acceptable way. Health workers still found it difficult to announce the disease to patients at the time of diagnosis and sometimes concealed certain information so as not to shock the patient. The expressions used to disclose the diagnosis played a role in patient’s acceptance of the condition and adherence to treatment. Prior research has underscored the importance of exercising caution when discussing sensitive topics, such as leprosy, within the context of a therapeutic relationship. [41]. Importantly, the care provided for those suffering from leprosy must be humane and contextualized in order to circumvent the exacerbation of stigma [42,43] and to facilitate the culturally appropriate utilization of biomedical treatments [44].

### Situating health seeking within a broader systemic perspective

Nevertheless, “looking at access to health care through an isolated ‘stigma’ lens can decontextualize the problem and lead to ineffective solutions” [45] and it remains important to consider all factors determining access to care and health seeking behavior beyond stigma and socio-cultural factors. In a country like Madagascar, active case detection is limited due to lack of logistical and financial resources at disposal of the National Leprosy Program (NLP), particularly in relation to the size and spread of the affected areas.

Several efforts have been made by the NLP and stakeholders to tackle leprosy, such as the introduction in 2015 of active case finding in some health centers combined with sensitization and patient care for leprosy and other common skin diseases [5,46]. During the PEOPLE study, very few refusals (0,1%) were reported for door-to-door screening. In terms of cases detected, a high prevalence of 35 per 10 000 was recorded during the first year of screening, followed by a low incidence rate of 0,57 per 10 000 in the subsequent two years [47]. This suggests that transmission is not rampant in our study context, yet passive case finding implies that a substantial number of cases go unnoticed. The study allowed for an increase in knowledge of health workers in the four district health centers and of community workers in the study sites. It has also enabled regular awareness-raising, active screening of patients and close monitoring of patients during their treatment, even if only for a limited number of years and in a restricted area.

However, while the fight against leprosy faces disease-specific challenges, it is also substantially constrained due to an underfunded, challenged and poor functioning health system more broadly, which remains very difficult to access in many rural parts of the country. As long as there is not an integrated approach allowing for people affected by different (skin) conditions to receive proper diagnosis and affordable treatment, even for simple dermatoses which is not currently the case, it will be difficult for people with minor symptoms, including early symptoms of leprosy, to consider the formal health system as a first option of care.

### Strengths and limitations of the study

To the best of our knowledge, this is the first scientific publication to describe community perceptions of leprosy in (one part of) Madagascar.

While we spoke with traditional healers, we were not able to observe consultations with traditional healers. Future studies could include such an observation component to better understand why people go to traditional healers and test an intervention engaging traditional healers to better orientate patients to the health centers when needed, as has been tried elsewhere.

This study was a qualitative study carried out in a limited number of places in one district of Madagascar, and therefore only discusses the context for the studied communities in this district. Given the island’s surface area and the fact that it is home to 18 ethnic groups, each with their own traditions and customs, leprosy perceptions may vary from one place to another. The results cannot be generalized.

## Conclusion

Our study demonstrates that understanding community perceptions of leprosy, its symptoms, its causes and dynamics, as well as the experience of stigma, are crucial when designing interventions that aim to increase the use of formal healthcare services, access and adherence to biomedical treatment for leprosy. While these findings echo already extensively described issues in the literature on leprosy, they highlight the remaining, actual need in some parts of Madagascar to improve access to formal health and skin care more broadly, to (re)train health staff on leprosy diagnosis and patient welcoming, to raise community awareness about warning symptoms for leprosy and to reduce stigma by communicating that leprosy can be healed if treated early-on.

## Data Availability

The data supporting the findings will not be made openly accessible due to confidentiality concerns as the dataset contains sensitive data and cannot be fully anonymized given the nature of the research. Data can, however, be made available after approval of a motivated and written request to the Institute of Tropical Medicine at.

## Acknowledgments

We are grateful to all the people we met during the fieldwork: the patients, community members, and healthcare workers, who accepted us in their communities and took their time to speak with us, the members of the PEOPLE project team, as well as all of those who kindly hosted us during our stay in the villages for the data collection.

## Notes

### Competing Interest Statement

The authors have declared no competing interest.

### Clinical Trial

This study was a socio-anthropological component of the PEOPLE project (Post-exposure prophylaxis in leprosy) and was registered with ClinicalTrials.gov: NCT03662022.

### Funding Statement

Yes

### Author Declarations

The research was approved by the Biomedical Research Ethics Committee of the Madagascar Ministry of Public Health (reference number: 032/MSANP/SG/-AGME /CNPV/CERBM). Address of the institution: Ministère de la Santé Publique – Ambohidahy BP.88 – ANTANANARIVO 101 The research was also approved by the Institutional Review Board of the Antwerp Institute of Tropical Medicine (reference number: 1255/18). Address of the institution: Nationalestraat 155, 2000 Antwerp, Belgium

